# Evolution of COVID-19 Health Disparities in Arizona

**DOI:** 10.1101/2021.12.27.21268462

**Authors:** Felix L. Shen, Jingmin Shu, Matthew Lee, Hyunsung Oh, Flavio Marsiglia, Ming Li, George Runger, Li Liu

**Affiliations:** Paradise Valley High School, Phoenix, AZ, 85032, USA; College of Health Solutions, Arizona State University, Phoenix, AZ, 85004, USA; Biodesign Institute, Arizona State University, Tempe, AZ, 85281, USA; School of Social Works, Arizona State University, Phoenix, 85006, AZ; Southwest Interdisciplinary Research Center, Watts College of Public Service and Community Solutions, Arizona State University, Phoenix, 85004, AZ; Phoenix Veterans’ Administration Health Care System, Phoenix, 85012, AZ; College of Medicine, University of Arizona, Phoenix, 85004, AZ

**Author notes:** Corresponding author: Li Liu.

**Keywords:** COVID-19, health disparity, population health

## Abstract

**Objective:** COVID-19 burdens are disproportionally high in underserved and vulnerable groups in Arizona. As the pandemic progresses, it is unclear if the disparities have evolved. In this study, we aim to elicit the dynamic landscape of COVID-19 disparities at the community level and identify newly emerged vulnerable subpopulations.

**Materials and Methods:** We compiled biweekly COVID-19 case counts of 274 zip code tabulation areas (ZCTAs) in Arizona from October 21, 2020, to November 25, 2021, during which the COVID-19 growth rate has changed significantly. Within each growth period, we detected health disparities by testing associations between the growth rate of COVID-19 cases in a ZCTA and the population composition of race/ethnicity, income, employment, and age. We then compared the associations between periods to discover temporal patterns of health disparities.

**Results:** High percentage of Latinx or Black residents, high poverty rate, and young median age were risk factors of high cumulative COVID-19 case counts in a ZCTA. However, the impact of these factors on the growth rate of new COVID-19 cases varied. While high percentage of Black residents and young median age remained as risk factors of fast COVID-19 growth rate, high poverty rate became a protective factor. The association between the percentage of Latinx residents and the COVID-19 growth rate converted from positive to negative during summer 2021. The unemployment rate emerged as a new risk factor of fast COVID-19 growth rate after September 2021. Based on these findings, we identified 37 ZCTAs that are highly vulnerable to fast escalation of COVID-19 cases.

**Discussion and Conclusion:** As the pandemic progresses, disadvantaged communities continue suffering from escalated risk of COVID-19 infection. But the vulnerabilities have evolved. While the disparities related to Latinx ethnicity improved gradually, those related to Black ethnicity and young communities aggravated. The struggle of financially disadvantaged communities continued, although the burden had shifted from those living under the poverty line to those with a high unemployment rate. It is necessary to adjust current resource allocations and design and deploy new interventions to address emerging needs.

## INTRODUCTION

COVID-19 disparities attributable to race/ethnicity and social determinants of health (SDoH) are prominent [1]. Ethnic minority groups and financially disadvantaged populations have experienced disproportionately high rates of COVID-19 incidence and mortality in the U.S.A. [2, 3]. Ecological studies examining various geographical boundaries such as state, county, and zip code tabulation areas (ZCTAs) highlighted substantial disparities in number of cases, testing, and fatality concentrated within historically underserved communities with disadvantaged SDoH [4-7]. In response to the early evidence of overwhelming disparities by race/ethnicity and key SDoH, various efforts have been established to mitigate social forces underlying these structural inequities during the pandemic [8-11].

Meanwhile, the pandemic has advanced through several waves. Given these developments, it is plausible that the disparity landscape has evolved. Understanding the changes of the disparities will inform population health task forces to better prioritize and serve the vulnerable groups. Unfortunately, current literature mainly contains cross-sectional studies capturing only static snapshots of the disparities, and updates of the COVID-19 disparities are surprisingly rare. To date, we found only one study that investigated the time-varying associations between COVID-19 incidence and community level risk factors during the early 7-month period (March to October 2020) in Massachusetts [12]. Overlooking the fluid nature of COVID-19 disparities impedes prompt responses to newly emerged needs.

To address this knowledge gap, we conducted a time-series study that tracked COVID-19 disparities over a 13-month period. We focused on communities in Arizona, home of more than seven million residents with diverse backgrounds. About 31.7% of Arizonans are Hispanic or Latino [13]. According to reports in 2020 [14, 15], this population had a 2 times higher risk of contracting the disease and an alarmingly 14.8-23.7 times higher risk of COVID-19 related death than non-Hispanic/Latino Whites in the state. Arizona also has a large Native American population (5.3%) including 22 Tribal Nations [13]. A study conducted in May 2020 [16], reported that Native Americans in Arizona had 3 times higher risk of COVID-19 infection and 4 times higher risk of death than the state average. National studies also reported COVID-19 disparities in other ethnic groups and low-income segments [1], which is expected in Arizona as well. Thus, it is necessary to examine how these features jointly influence COVID-19 prevalence at different stages of the pandemic.

Because individual-level data are not readily available, we used COVID-19 case counts aggregated by zip code areas in Arizona, which is updated daily on the Arizona Department of Health Services (ADHS) Dashboard [17]. Using this public resource, we collected time series data and tested associations of COVID-19 prevalence and growth rate with zip-code level demographic characteristics. We confirmed the initial COVID-19 disparities reported in previous studies and elicited the temporal changes. We discovered that disparities related to Hispanic or Latino ethnicity gradually improved while disparities related to Black or African American ethnicity worsened. Meanwhile, the vulnerable groups based on income shifted from populations living under the poverty line to populations of unemployment. The high prevalence of COVID-19 in young communities persisted. Based on these patterns, we identified 37 zip code areas with high risk of disproportionately fast growth of COVID-19 cases and 22 zip code areas with low risk.

## MATERIALS AND METHODS

### Datasets

As recorded in the most recent (2019) U.S. Census [13], Arizona has 387 active zip codes that have more than 100 residents. We collected demographic and COVID-19 data of these zip code areas from public databases.

#### Demographic data

For each active zip code area in Arizona, we queried the US 2019 Census Database [13] to retrieve 12 demographic features, including total population, population density, and population compositions on sex (ratio of male and female), age (median age, Seniors over 65 years old), ethnicity (White non-Hispanic or Latino, Hispanic or Latino, American Indian, Black, and Asian), and household financial status (median income, poverty rate, and unemployment rate).

#### COVID-19 data

We downloaded case counts aggregated by zip code from the ADHS Dashboard website [17] every two weeks over a 13-month period from October 21, 2020, to November 25, 2021. Case counts fewer than 10 were suppressed due to HIPAA compliance and were substituted as 5. Because ADHS Dashboard suppresses COVID-19 data for Indian tribes, we removed zip code areas that overlap with Indian tribes to avoid biases. The remaining 274 zip code areas were subject to statistical analyses.

### Estimating COVID-19 prevalence and growth rate

#### Detecting growth periods at the State level

The growth rate of COVID-19 cases in Arizona was not constant, experiencing waves of acceleration and deceleration. To identify distinct growth periods, we performed segmented regression analysis using the R/strucchange package [18]. We first prepared the time series data by aggregating new case counts over all Arizona zip code areas on a biweekly basis, then applying log transformation to account for the exponential growth. We next conducted the Andrew’s supF test followed by the Chow test to search for significant structural changes in the linear regression models fit to the time series data. Breakpoints with a Chow test p-value <0.05 were boundaries of growth periods.

#### Quantifying COVID-19 prevalence and growth rate at the zip code level

We denote the total case count in a zip code area *z* at a time point *t* as *c*_*z,t*_. Because the population size *s*_*z*_ of a zip code area affects case counts, we computed a normalized 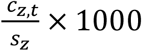 value representing prevalence per 1,000 residents (cases / 1K).

To estimate the COVID-19 growth rate in a zip code area *z* during a time interval *τ*, we used counts of new cases Δ*c*_*z,τ*_ and computed a normalized 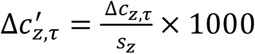 value representing new cases per 1,000 residents. We further applied log transformation and defined the growth rate as 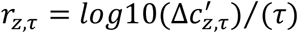, where (*τ*) is the length of the time interval in weeks. We used this approach to estimate the growth rate for every zip code area during every biweekly interval and during every growth period.

### Discovering COVID-19 health disparities

#### Identifying demographic features associated with COVID-19 prevalence or growth rate

To find which demographic features were associated with COVID-19 growth rate at which time intervals, we performed bidirectional stepwise feature selection. Given a time interval *τ*, we tested the relationship between the growth rate of COVID-19 cases in a zip code area and a set of demographic features *D*_*τ*_ using a multiple linear regression model

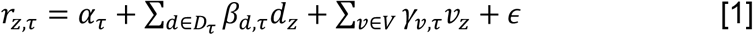

where *α*_*τ*_ is the intercept, *β*_*d,τ*_ is the coefficient of a demographic feature, *d* ∈ *D*_*τ*_, *γ*_*ν,τ*_ is the coefficient of a covariate *ν* ∈ *V* that includes population density as a proxy of transmission probability and median age, and *ϵ* is Gaussian-distributed error. In the forward stepwise selection stage, we progressively expanded *D*_*τ*_ by adding a feature with the lowest and significant p-value (<0.05) associated with the corresponding *β*_*d,τ*_ until all features *d* ∉ *D*_*τ*_ had p-value>0.05. In the backward stepwise selection stage, we progressively shrank *D*_*τ*_ by removing a feature *d* ∈ *D*_*τ*_ with p-value >0.05 until all features *d* ∈ *D*_*τ*_ had p-values <0.05. The final set of features *D*_*τ*_ was jointly associated with COVID-19 growth rate at the time interval *τ*. We performed this analysis for every biweekly interval and for every growth period.

By replacing the response variable in equation (1) with log transformed COVID-19 prevalence, we identified demographic features associated with COVID-19 prevalence at every time point.

#### Examining the dynamics of COVID-19 health disparities

For each demographic feature *d*, we estimated its impact on the growth rate of COVID-19 cases during a time interval *τ* using multiple linear regression. For a feature *d* ∈ {*D*_*τ*_, *V*}, its impact was the corresponding coefficient estimated in equation (1). For a feature *d* ∉ {*D*_*τ*_, *V*}, we included it as an additional term to equation (1) such that *r*_*z,τ*_ = *α*_*τ*_ + ∑*d*∈*D*_*τ*_ *β*_*d,τ*_*d*_*z*_ + ∑*ν*∈*V γ*_*ν,τ*_*ν*_*z*_ + *ω*_*τ*_*d* + *ϵ*, where *ω*_*τ*_ represented its impact. To correct for multiple comparisons, we adjusted the nominal p-value associated with each coefficient using the Benjamini-Hochberg method [19] and computed false discovery rate (FDR). FDR <0.1 indicated a significant association between a feature and the growth rate of COVID-19 cases, i.e., significant health disparity. To examine the change of health disparities, we plotted the coefficients of each demographic feature over the entire study period.

We performed these analyses in R and Python. Data sets and source code are available at https://github.com/liliulab/COVID-19-Disparities.

### RESULTS

### Surges and declines of COVID-19 cases

The segmented linear regression model revealed 6 distinct periods between which the growth rate of COVID-19 cases at the State level changed significantly (all Chow test p-values < 0.05, **Fig. 1A**). The Surge-A period and the Decline-A period marked a wave of COVID-19 new cases between October 21, 2020, and March 15, 2021. It was followed by a four-month Valley period during which the COVID-19 growth rate remained low till July 7, 2022. After that, it spiked again during the Surge-B period, dropped slightly during the Decline-B period, and rose again during the Surge-C period that persisted till the end of our study (November 25, 2021). The growth rates at the zip code level were significantly different between these periods (all t test p-values <10^−7^, **Fig. 1B**).

**Figure 1.**
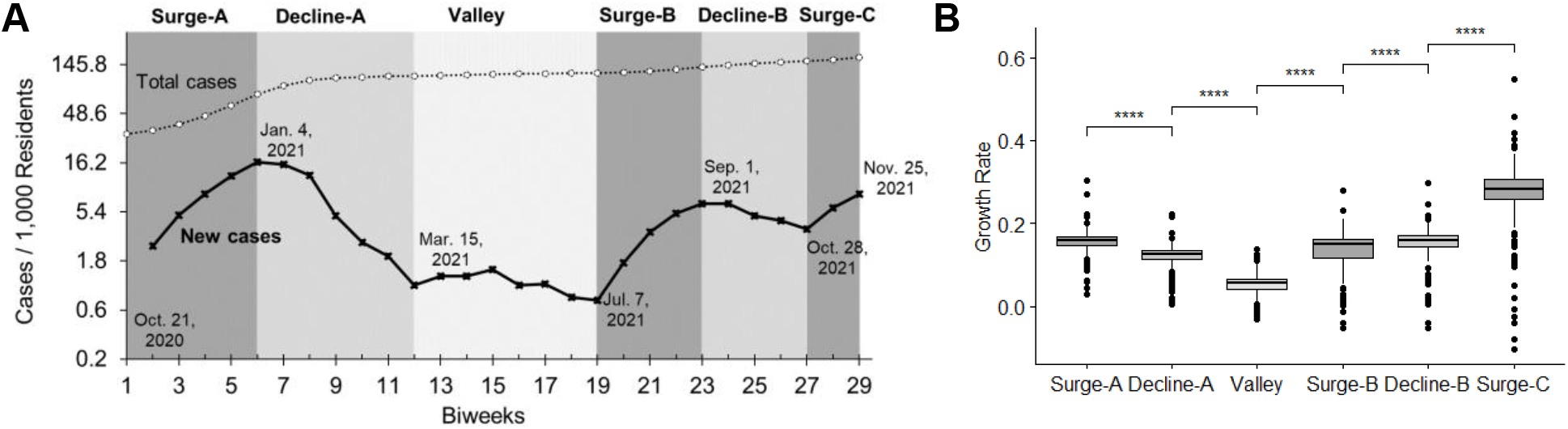
Time intervals with distinct growth rates of COVID-19 prevalence. (**A**) Biweekly counts of new cases and cumulative cases in Arizona. Distinct periods are shaded with different gray colors. Dates marking the boundaries are shown. (**B**) Boxplots of growth rates of COVID-19 cases at the zip code level in each growth period. Asterisks indicate significant difference of growth rates between adjacent periods.

### Evolution of COVID-19 disparities

Using bidirectional stepwise feature selection, we found different demographic features associated with the grow rates of COVID-19 cases during different time intervals (**Supplementary Table 1 and 2**). The associations varied as the pandemic progressed, shifting between significant and insignificant impact, and sometimes showing opposite effects. Such temporal changes implied evolution of COVID-19 disparities.

#### Ethnicity-based disparities

Hispanic or Latino is the largest ethnic minority in Arizona, representing 31.7% of the total population. In 20.8 % of the zip code areas we examined, Hispanic/Latino is the dominant ethnicity, surpassing non-Hispanic/Latino White (**Fig. 2A**). At the beginning of this study on October 21 2020, our analysis showed COVID-19 prevalence in a zip code area was positively associated with the percent of Hispanic/Latino population (P=2×10^−12^, **Fig. 2B**), consistent with previous reports [14, 15]. Furthermore, the growth rate of COVID-19 cases increased with the percentage of Hispanic or Latino population for about 6 months spanning the Surge-A period and the Decline-A period (all FDR<0.002, respectively, **Fig. 2C**), implying aggravation of the disparate impact. Fortunately, as the pandemic entered the Valley period, the association between the growth rate of new cases and the percentage of Hispanic or Latino population disappeared (P=0.38), and then became negative during the following wave including the Surge-B and Decline-B periods (all FDR<0.02). These observations suggested that the disparity associated with Hispanic/Latino ethnicity has improved since March 2021. However, it is worth noting that the positive association reappeared during the Surge-C period, although it was not statistically significant (P=0.58).

**Figure 2.**
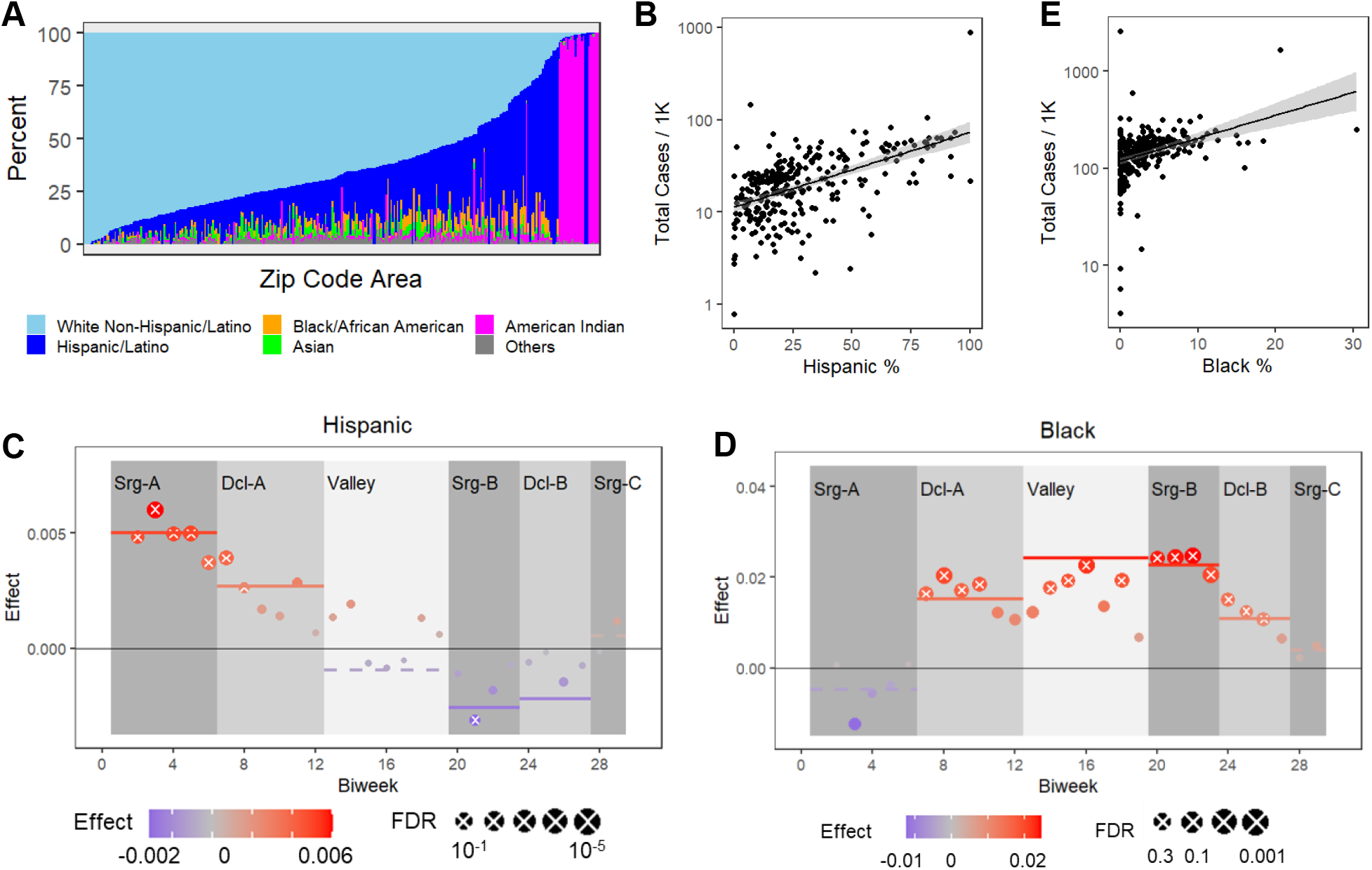
Impact of Ethnicity. (**A**) The stacked bar plot shows the percentages of various ethnic populations in the 274 zip code areas. (**B**) The scatter plot shows the positive correlation between the percentage of Hispanic or Latino population and the COVID-19 prevalence at the beginning of the study on Oct. 21, 2020. (**C, D**) The effect of ethnicity on the growth rate of COVID-19 cases are shown for every biweekly interval (circles) and for each growth period (lines). Crosses and solid lines indicate statistically significant effects with FDR<0.1. Effects are measured by coefficients corresponding to the percentage of Hispanic or Latino population (C) or coefficients corresponding to the percentage of Black or African American population. (**E**) The scatter plot shows the positive correlation between the percentage of Black or African American population and the COVID-19 prevalence at the end of the study on Nov. 25, 2021.

American Indian is the second largest ethnic minority in Arizona, representing 5.3% of the total population. Because COVID-19 case counts are suppressed for American Indian tribes, our data unfortunately covered only non-tribe zip code areas where merely 1.9% of population is American Indian. Using this data set, we did not observe significant associations between the percentage of American Indian population and the prevalence or the growth rate of COVID-19 during any period.

Black or African American is the third largest ethnic minority in Arizona, representing 5.2% of the total population. At the beginning of the study, the positive association between the COVID-19 prevalence and the percentage of Black or African American population was at the borderline (P=0.052). However, zip code areas with a high percentage of Black or African American residents experienced a fast growth of new cases during the 10 months spanning the Decline-A, Valley, Surge-B, and Decline-B periods (all FDR <0.07, **Fig. 2D**). Consequently, at the end of the study on November 25, 2021, the association between the percentage of Black or African American and the COVID-19 prevalence had become positive and significant (P=0.015, **Fig. 2E**). These observations suggested that the disparity associated with the Black or African American ethnicity has worsened over the time.

For the other ethnicities including non-Hispanic or Latino White and Asian that respectively represents 54.1% and 3.7% of Arizona populations, we did not detect significant associations with the COVID-19 prevalence or the growth rates at most of the time intervals. The only exception was the during the Valley period where the Asian ethnicity showed a transient positive association with the growth rate of COVID-19 cases, which quickly disappeared afterwards.

#### Income-based disparities

We collected three financial features, including the median household income, poverty rate, and unemployment rate. Because ethnic minority groups are financially disadvantaged, we examined how the financial status of a zip code area influenced the COVID-19 growth rate after accounting for the percentage of Hispanic or Latino population and the percentage of Black or African American population.

The median household income and the poverty rate in a zip code area are negatively correlated (Pearson correlation coefficient PCC= –0.70, P=10^−40^). Surprisingly, our multiple regression analysis showed that the COVID-19 prevalence increased with the poverty rate as well as the median household income at the beginning of the study (P=1×10^−2^ and 9×10^−6^, respectively, **Fig. 3A-B**). However, their associations with the growth rate differed. While fewer new cases were reported in zip code areas with a high poverty rate, more new cases were reported in areas with a high median income during various periods (**Fig. 3B-D**). Consequently, at the end of the study, the COVID-19 prevalence was no longer associated with the poverty rate (P=0.25) but remained positively associated with the median household income (P=0.009).

**Figure 3.**
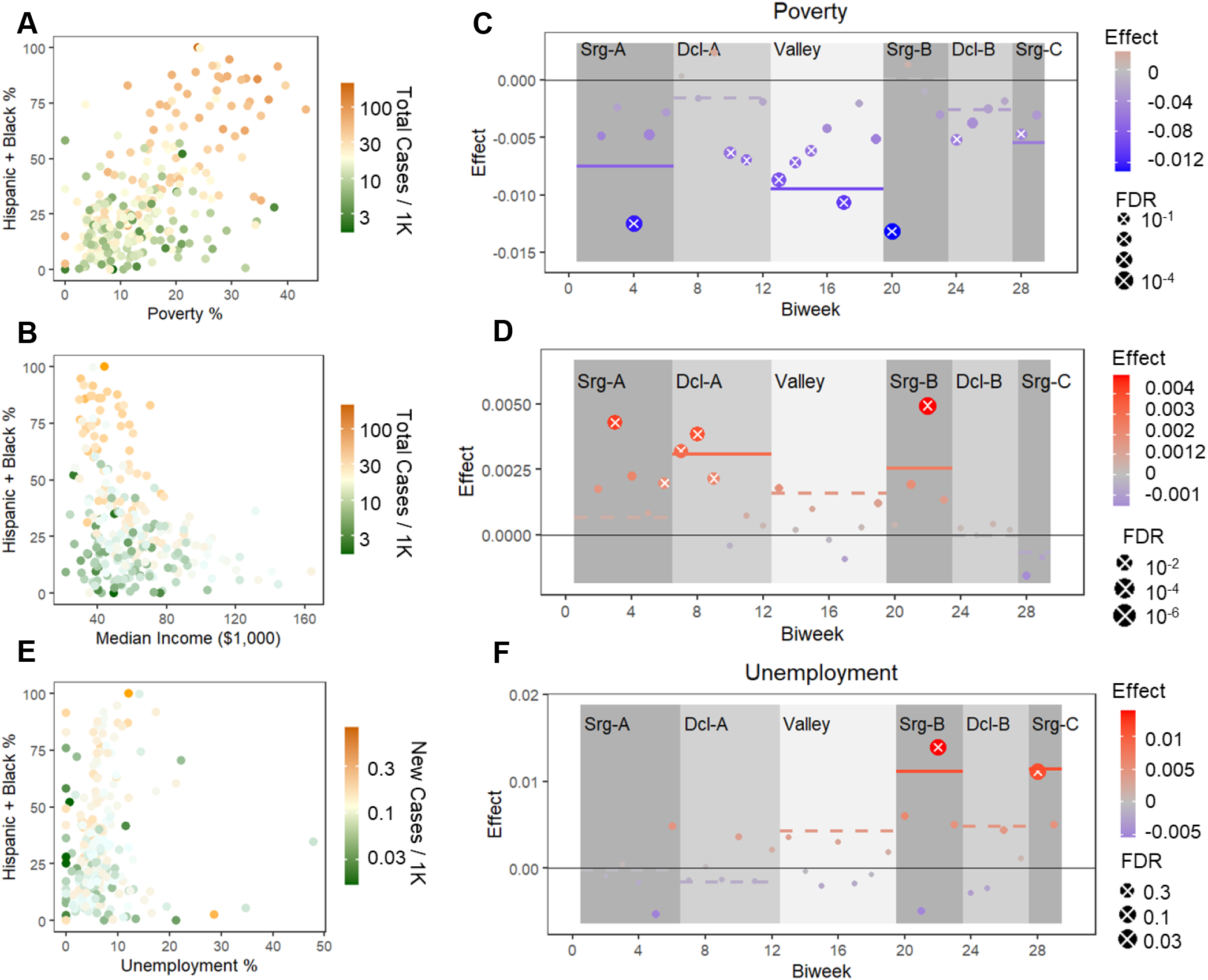
Impact of financial status. (A) The scatter plot with heatmap colors shows the COVID-19 prevalence at the beginning of the study increased with the poverty rate and the percentage of vulnerable ethnicities including Hispanic or Latino and Black or African American. (B) The scatter plot with heatmap colors shows the COVID-19 prevalence at the beginning of the study decreased with the median income. (E) The scatter plot with heatmap colors shows cumulative new cases over the 13-month study period increased with the unemployment rate and the percentage of vulnerable ethnicities. (C, D, F) The effect of financial status on the growth rate of COVID-19 cases are shown for every biweekly interval (circles) and for each growth period (lines). Crosses and solid lines indicate statistically significant effects with FDR<0.1. Effects are measured by coefficients corresponding to the poverty rate (C), coefficients corresponding to household median income (D), or coefficients corresponding to the unemployment rate (F).

The unemployment rate emerged as a new potential factor of COVID-19 disparities. At the beginning of this study, it was not associated the COVID-19 prevalence (P=0.77). However, as the pandemic progressed into the Surge-B and Surge-C periods, the number of new cases increased unproportionally fast in zip code areas with a high unemployment rate (**Fig. 3E**). At the end of the study, the new case count aggregated over the 13 months has become positively associated with the unemployment rate (P=0.046, **Fig. 3F**).

#### Persistent impact of age

The median age of zip code areas in Arizona has a bimodal distribution (**Fig. 4A**). The first peak at 36 years old (yo) is close to the State median age (37.9 yo). The second peak at 62 yo reflects the large number of senior communities in Arizona.

**Figure 4.**
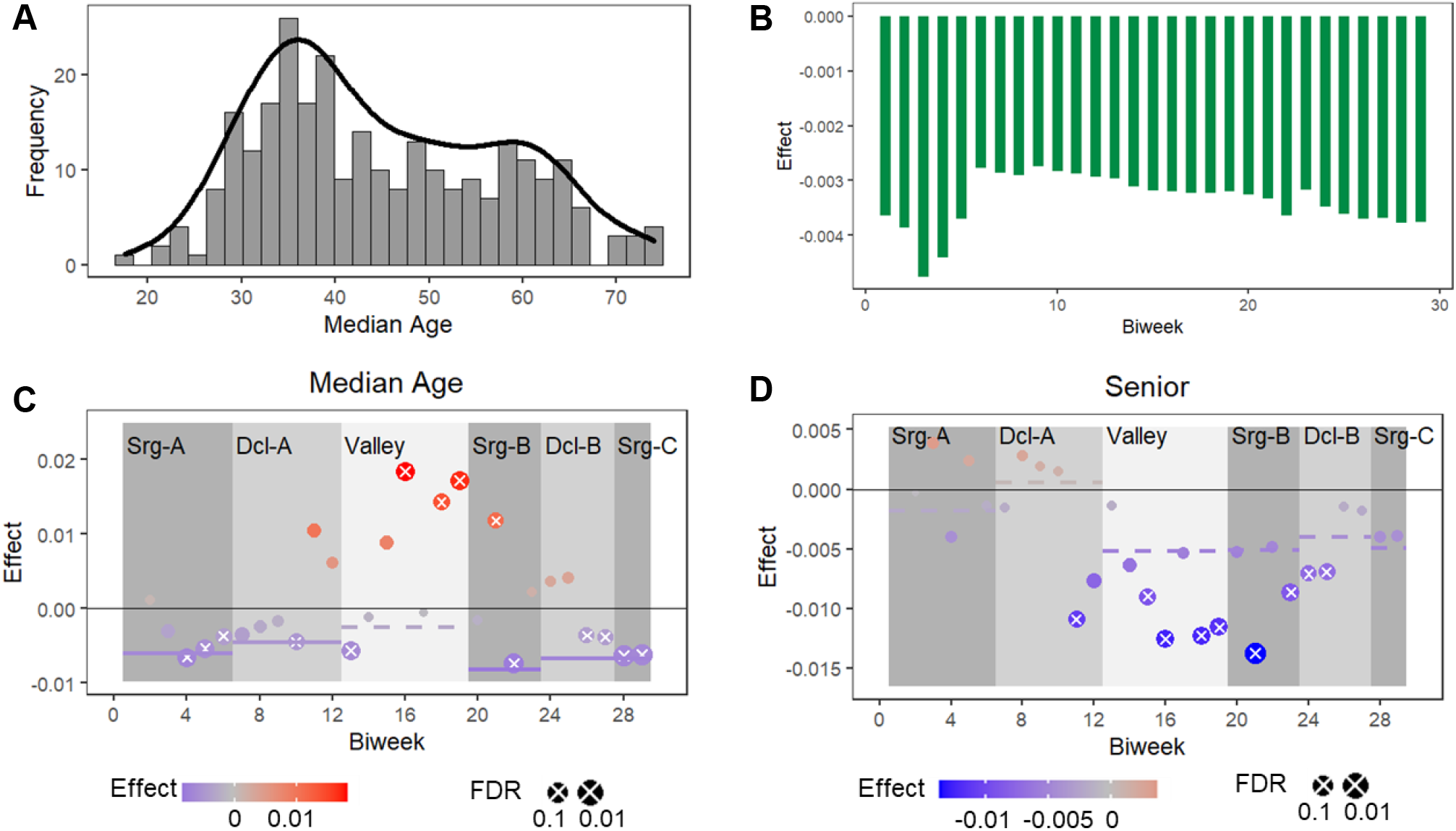
Impact of age. (**A**) The histogram shows a bimodal distribution of median age in Arizona zip code areas. (**B**) The bar plot shows the effect of median age on COVID-19 prevalence was negative at every time point. (**C, D**) The effect of age on the growth rate of COVID-19 cases are shown for every biweekly interval (circles) and for each growth period (lines). Crosses and solid lines indicate statistically significant effects with FDR<0.1. Effects are measured by coefficients corresponding to the median age (C), or coefficients corresponding to the percentage of senior residents (>65 years old).

We found that COVID-19 impacted younger communities greater than it did older communities. The median age in a zip code area was negatively associated with the COVID-19 prevalence for all biweekly time points we examined (all FDR<0.02, **Fig. 4B**). It was also negatively associated with the growth rate during most time intervals (**Fig. 4C**). The only exception was in the Valley period where the percentage of senior residents (>65 yo) instead of the median age were negatively associated with the growth rate. The indications remained the same.

### Risk stratification

Our analysis showed that high percentage of Hispanic or Latino population, high percentage of Black or African American population, high poverty rate, high unemployment rate, and low median age are risk factors of fast COVID-19 growth. Using the median value of each risk factor as the cutoff, we classified 37 zip code areas meeting all risk criteria as high risk, 22 zip code areas meeting no risk criteria as low risk, and the remaining areas as median risk (**Fig. 5A**). The high-risk areas are clustered in the central Phoenix metropolitan area and the central Tucson area. Interestingly, many high-risk areas are adjacent to low-risk areas (**Fig. 5B-C**). We confirmed that the high-risk group and the low-risk group had significantly different growth rates during all periods (all t test P <0.002, **Fig. 5D**). Therefore, ethnicity and SDoH, instead of geographics, predispose these populations to different levels of vulnerability.

**Figure 5.**
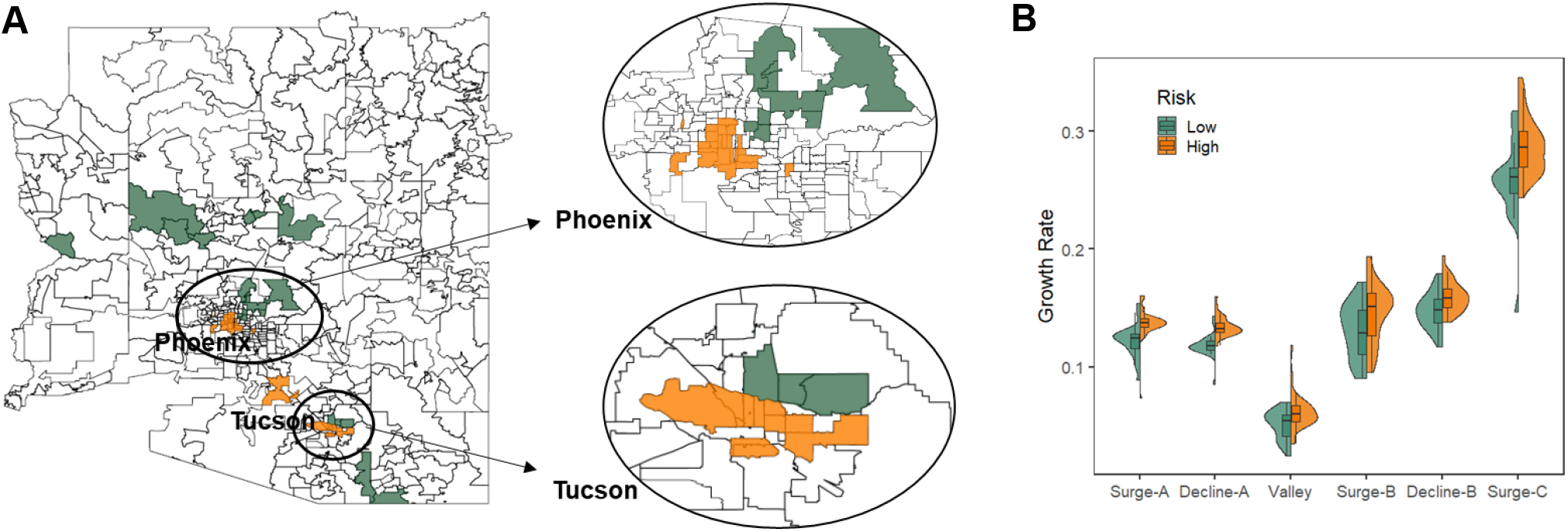
Risk groups. (**A**) The map of Arizona zip code areas shows geographic locations of the high-risk areas (orange) and low-risk areas (green). (**B**) The violin plot shows distributions of growth rates of the high-risk and low-risk groups during each growth period.

## DISCUSSION

COVID-19 is placing unprecedent challenges on our communities, especially on socially and economically disadvantaged groups [1]. While existing studies captured only snapshots of the situations using cross-sectional data, we tracked structural health disparities in Arizona over 13 months and discovered that COVID-19 vulnerability related to ethnicity and financial status has evolved.

At the beginning of the study, zip code areas with a high percentage of Hispanic or Latino population and a high poverty rate experienced disproportionately high COVID-19 prevalence and fast growth. This situation improved over time, with the trend reversing after July 2021, and fewer new cases were reported in these areas than the state average. This change may reflect the outcomes of the multiple programs that have been established in Arizona to support Hispanic or Latino communities and low-income populations during the pandemic [8-11]. Acquired immunity through previous infection or vaccination may also contribute to the slow-down. However, we observed signs of reappearance of the disparities during the Surge-C period, suggesting needs for augmentation of the support programs.

Meanwhile, COVID-19 cases grew very fast in areas with a high percentage of Black or African American populations and a high unemployment rate. At the end of our study, these two features have emerged as significant risk factors, suggesting inadequate public health responses and urgent needs of support to these communities to ameliorate the impact.

Arizona has a large aging population, with senior residents comprising the majority in many zip codes. Our analysis showed that COVID-19 growth rates are relatively low in these areas, suggesting these populations are relatively well supported and protected.

Our study has several limitations. First, because individual-level data are not readily available, we used zip code level data. Thus, the identified vulnerable groups and their evolving patterns had restricted precision. However, the publicly available zip code level data allows us to track COVID-19 disparities easily and continuously throughout the present and into the future. Second, we did not include COVID-19 vaccination data in our analysis, although vaccination rates may have significant associations with COVID-19 prevalence and growth rates. This was again due to data availability. Third, the pandemic has disruptive impact on the financial status of many families [20, 21]. Such changes were not incorporated in our analyses because the population composition data were based on the 2019 US Census.

Although our study is restricted to Arizona data, we expect that COVID-19 disparities in other regions may have also evolved. To better support disadvantaged populations, we need to monitor the progression and adjust resource allocations accordingly.

## CONCLUSION

As the COVID-19 pandemic is dynamic, so are the structural health disparities, which have intensified during this timeframe. In Arizona, we continue to support Hispanic and Latino communities and low-income populations to fight the disease. However, actions are urgently required to enhance the situations in Black or African American populations and unemployed populations.

## Supporting information

Association test results for each variable at each biweekly time point

Association test results for each variable at each growth period

## Data Availability

All data produced are available online at https://github.com/liliulab/COVID-19-Disparities

## CONTRIBUTION STATEMENTS

L.L., H.O., F.M., and G.R. conceived of the presented idea. F.L.S., J.S., M.L, and L.L. conducted the analysis. All authors contributed to the manuscript.

## ACKNOLWEDGEMENT

This study is supported by the NIH grant 5U54MD002316-14.

